# Differential Predictability of Preterm Birth Types: Strong Signals for Indicated Cases versus Limited Success in Spontaneous Preterm Birth

**DOI:** 10.1101/2025.07.09.25329712

**Authors:** Yun Chao Lin, Andrea Clark-Sevilla, Mahdi A. Loodaricheh, Itsik Pe’er, Anita Raja, Ronald Wapner, Ansaf Salleb-Aouissi

## Abstract

**Background:** Preterm birth, defined as birth occurring before 37 weeks of gestation, poses a significant and enduring public health challenge, with substantial emotional and financial burdens on families and society. To identify preterm births early in pregnancy, we investigated the predictive ability of machine learning models in a nulliparous (first-time pregnancy) study cohort. Preterm births are categorized into two major types: indicated preterm birth, which occurs due to medical conditions such as preeclampsia or other maternal/fetal complications requiring early delivery, and spontaneous preterm birth, which involves the natural onset of preterm labor. Our research aims to develop predictive tools that could enable earlier intervention and improved outcomes for these vulnerable pregnancies.

**Methods:** Our study analyzed the Nulliparous Pregnancy Outcomes Study: Monitoring Mothers-to-be cohort (nu-MoM2b), comprising data from eight clinical sites throughout the United States from October 2010 to May 2014, including treatment, psychological, physiological, medical history, demographic, ultrasound, activity, toxicology, family history, pre-pregnancy diet, and genetic race. We distinguished between spontaneous and indicated preterm births to develop targeted predictive models for each subtype. We also used a novel approach to predict preterm birth called learning with privileged information, information available during training but often inaccessible during evaluation. Specifically, the set of privileged information that we utilized for PTB prediction includes the occurrence of adverse pregnancy outcomes (APOs), after-delivery physiology information, and maternal outcomes. We developed an enhanced model, XGBoost+, which incorporates this privileged information to improve predictive performance compared to traditional machine learning approaches.

**Results:** We selected XGBoost as our base model due to its robust performance with tabular data and its ensemble approach that effectively mitigates overfitting while capturing complex relationships between clinical variables, making it particularly well-suited for the heterogeneous risk factors associated with preterm birth prediction. XGboost-based models achieved higher AUC against all other models, including decision tree, random forest, logistic regression, and SVM for all visits. Our XGboost+ model, utilizing privileged information, achieved an AUC of 0.72. Analyzing the subcategories of preterm birth, XGboost+ achieved similar performance with XGboost for spontaneous preterm birth (0.68 AUC versus 0.67 AUC), but improvements were more significant for indicated preterm birth (0.78 versus 0.74). These results demonstrate the benefits of how information that is not typically utilized in traditional machine learning models can help build better models.

**Conclusion:** Our extensive analysis of this comprehensive set of risk factors revealed preterm birth as a multifaceted issue, with different risk factors associated with two subcategories of preterm birth - spontaneous and indicated. Notably, we achieved significant success in predicting indicated preterm birth, demonstrating strong predictive performance (AUC 0.78) using our XGBoost+ model. This finding represents an important advancement, as indicated preterm birth is influenced mainly by conditions related to hypertension and preeclampsia, which our model effectively captured. While spontaneous preterm birth remains challenging to predict with clinical data alone, especially in early pregnancy, our research successfully differentiates between these subtypes and provides a valuable predictive tool for indicated preterm birth. The complexity of spontaneous preterm birth suggests that future research should focus on gathering more proximal biological data, including vaginal microbiota or raw cervical images, to complement our successful approach for indicated preterm birth prediction.

## Introduction

Premature or preterm birth (PTB) is defined as birth before 37 weeks of gestation. It is a long-lasting major public health problem with emotional and financial consequences for families and society.^1^ PTB is the leading cause of long-term mortality and disability among newborns. These range from visual/hearing impairment, cerebral palsy, and mental retardation to an increased risk of cardiovascular disease, hypertension, and diabetes later in life.^2^ Furthermore, more than 26 billion dollars are spent annually on the delivery and care of approximately 12% babies born prematurely in the United States.^3^ A crucial challenge is to identify women who are at the highest risk for extremely preterm birth to facilitate interventions. Equally important would be identifying women at the lowest risk to avoid unnecessary and costly interventions. Despite decades of research and efforts, preterm birth rates worldwide are still alarming.^4,5^

Understanding the heterogeneity of preterm birth is essential for developing effective prediction models. Approximately 30% of preterm deliveries are indicated based on maternal or fetal indications, such as preeclampsia or other complications requiring early delivery. The remaining 70%, classified as spontaneous PTB (sPTB), occurs after the onset of spontaneous preterm labor, prelabor premature rupture of the membranes (pPROM), and cervical insufficiency.^6^ Spontaneous preterm labor is a particularly heterogeneous condition, the final common product of numerous biologic pathways that include immune, inflammatory, neuro-endocrine, and vascular processes.^3^ This distinction between indicated and spontaneous preterm birth, though critically important for prediction and intervention, has been inadequately addressed in previous research.

Currently, the standard of care for preventing preterm birth (PTB) has evolved significantly. Previously, treatment with 17-alpha hydroxyprogesterone caproate (17P or 17-OHPC) via intramuscular injections was widely recommended for women with a history of preterm delivery.^7,8^ However, in 2020, the FDA requested withdrawal of Makena (the branded 17P treatment) due to a confirmatory trial failing to demonstrate significant effectiveness in preventing preterm births. The current approach now primarily uses vaginal progesterone, which is more targeted and focuses on specific risk factors such as short cervical length. This change in treatment protocol continues to focus mainly on women with a history of preterm delivery, a risk factor with recurrence rates as high as 50% depending on the number and gestational age of previous deliveries.^6,9^ Critically, this approach still largely excludes nulliparous women (first time mothers), who represent approximately 40% of all pregnancies and have no history of pregnancy to guide risk assessment.

Epidemiological research has identified numerous other risk factors for preterm birth. In the United States, women classified as black, African-American, and Afro-Caribbean are at increased risk.^6^ Additional risk factors include low socioeconomic status, extreme maternal age, single marital status;^10,11^ low pre-pregnancy body mass index;^12^ high-risk behaviors during pregnancy; psychological factors such as depression^13^ and high stress levels;^14,15^ obstetric conditions including intrauterine infection, uteroplacental ischemia, hemorrhage, placental vascular lesions, uterine overdistention, and cervical insufficiency;^16,17,18,19^ closely spaced gestations;^20^ multiple gestations;^6^ and assisted reproductive technologies.^21,22^ Environmental factors, particularly tobacco smoke exposure,^23,24^ and genetic predisposition^25,26,27^ have also been implicated. We have conducted extensive analysis of risk factors related to preterm birth; details of related risk factors on preterm risk factors and their odd ratio are available in our prior scoping review of preterm birth risk factors.^28^ Furthermore, our prior work extends to analyzing preterm risk factors in national datasets, such as birth data from the Center for Disease Control (CDC).^5^

Clinical assessment for preterm birth risk has typically relied on several key factors. Research has shown^29^ that the odds ratio of sPTB was highest for fetal fibronectin, followed by a short cervix and a history of previous PTB. Systematic reviews^30,31^ have demonstrated that cervical length (threshold of 25 mm) measured by transvaginal ultrasound in high-risk women is highly predictive of spontaneous preterm birth before 35 weeks. However, the reliance on previous preterm birth history as a predictor leaves a significant gap in risk assessment for nulliparous women.

Previous attempts to predict preterm birth have been limited by several factors: study populations that were small or poorly characterized; insufficient attention to the critical distinction between spontaneous and indicated preterm birth; limited timing and variety of biological samples; and inadequate consideration of the multifactorial, heterogeneous nature of PTB. These limitations have hindered the development of effective prediction models, particularly for nulliparous women who lack prior pregnancy history.

To address these critical gaps, our study leverages the nuMoM2b dataset, an unparalleled pregnancy cohort of nulliparous women prospectively enrolled specifically for preterm birth prediction. We employ machine learning approaches to develop models capable of distinguishing between different types of preterm birth, with a particular focus on the fundamental differences between spontaneous and indicated preterm birth subtypes. Our research introduces a novel methodological approach called learning with privileged information (XGBoost+), which utilizes data available during model training but typically inaccessible during clinical evaluation. This innovative methodology allows us to investigate the predictability of both spontaneous and indicated preterm birth separately.

Our findings reveal important differences in the underlying mechanisms and predictability of these two preterm birth subtypes. We demonstrate that indicated preterm birth shows significantly higher predictability through its association with specific clinical conditions such as hypertension and preeclampsia, while spontaneous preterm birth presents greater prediction challenges. This work represents a significant step toward developing targeted prediction models that could enable more effective intervention strategies for women at risk, particularly nulliparous women who are currently excluded from standard preventive treatments.

### Related Work

The quest to predict preterm birth has a long history, beginning with early scoring systems. Papiernik proposed in the late 1960s an empirical method for estimating premature delivery risk called “The Coefficient of Premature Delivery Risk (CPDR)”.^32^ This approach grouped maternal characteristics into four categories (social status, obstetric history, work conditions, and pregnancy characteristics) in a two-dimensional table, assigning points to each characteristic according to importance. The Papiernik risk table was later modified for the risk of preterm delivery (RPD) system proposed by.^33^ However, a further assessment of Creasy’s table^34^ in different populations demonstrated a low predictive performance.

Early statistical approaches showed limited success.^35^ proposed a graded risk system in the NICHD MFMU preterm prediction study. However, their results of their multivariate logistic regression were modest: sensitivity of only 24.2% and 18.2%, with specificity of 28.6% and 33.3% for nulliparous and multiparous women, respectively. While this study represented a first step toward combining multiple risk factors, it highlighted the need for more sophisticated techniques to identify at-risk women better. Despite these efforts, no widely tested risk scoring/prediction system effectively combines the multifactorial aspects of PTB.^36^ The NIH^2^ and NICHD^37,38^ have recognized this gap and the need to discover the multifactorial etiologies of PTB for accurate prediction.

The application of machine learning to preterm birth prediction represents a more recent development. Goodwin et al. explored data mining techniques for this purpose, initially showing the feasibility of generating expert system rules for predicting preterm delivery.^39^ In subsequent work,^40^ they analyzed data from Duke University Medical Center (19,970 patient records with 1,622 variables) and identified seven demographic variables (maternal age, marital status, race, education, patient insurance category, county, and religion) as particularly important predictors. However, concerns persist about the demographic representativeness of their sample and the reproducibility of their results. Courtney et al.^41^ later demonstrated that this demographic model could generalize to broader populations, though with only modest accuracy.

A critical gap in previous approaches is their limited applicability to nulliparous women (first-time mothers). The current standard of care administers preventive treatment only to women with prior preterm births,^42^ excluding nulliparous women, who represent approximately 40% of pregnant women. Our preliminary work developed predictive scores for PTB based on non-genetic maternal attributes,^43,44,45^ highlighting the need for models that can identify at-risk nulliparous patients without relying on prior birth history.

Our current research extends previous work in three important ways. First, we specifically target nulliparous women, addressing a critical gap in clinical practice where there are no validated prediction tools for this population. Second, we employ advanced machine learning techniques, particularly our novel XGBoost+ approach, which incorporates privileged information that goes beyond traditional methods to leverage data available during training but inaccessible during clinical evaluation. Third, we explicitly distinguish between the spontaneous and indicated subtypes of preterm birth, which previous prediction efforts have often failed to differentiate despite their distinct etiologies.

We used the nuMoM2b data set (Nulliparous Pregnancy Outcomes Study Monitoring Mothers-to-be), which focuses on pregnancy outcomes for 10,038 nulliparous mothers with singleton gestations made available by the NIH-NICHD specifically for this challenging cohort.^46^ This comprehensive data set allows us to develop a dynamic risk prediction system that addresses the nulliparity gap and the differential predictability of preterm birth subtypes, potentially benefiting up to 200,000 pregnancies annually in the US alone through early identification and intervention.

## Methods

### Study design

In this study, we utilized the Nulliparous Pregnancy Outcomes Study: Monitoring Mothers-to-be (nuMoM2b) cohort,^46^ a meticulous, prospective cohort study specifically designed to investigate pregnancy outcomes in nulliparous women. This diverse cohort encompasses participants from eight clinical centers across the United States (Case Western Reserve University, Columbia University, Indiana University, University of Pittsburgh, Northwestern University, University of California at Irvine, University of Pennsylvania, and University of Utah). A total of 10,038 nulliparous women with singleton gestations were enrolled between September 30, 2010, and September 23, 2013, and followed prospectively from early pregnancy through delivery.

The nuMoM2b cohort offers several key advantages for studying preterm birth, particularly the ability to distinguish between indicated and spontaneous preterm births. Participants’ demographic characteristics, clinical measurements (e.g., blood pressure, cervical length), sonographic findings (e.g., uterine artery Doppler waveforms), exposures (e.g., diet, stress, toxicology), and pregnancy outcomes were collected with high precision and standardization. All data were collected by research personnel who underwent centralized training and certification for this cohort, ensuring consistency and reliability across all sites.

Structured data collection occurred at three time points during gestation, corresponding with routine clinical visits at each trimester: study visit 1 (6^0^ through 13^6^ weeks gestation), study visit 2 (16^0^ through 21^6^ weeks gestation), and study visit 3 (22^0^ through 29^6^ weeks gestation).

During study visit 1, comprehensive baseline data were collected, including blood pressure, anthropometric measurements (height, weight, waist, hip, and neck circumferences), and detailed interviews covering demographic characteristics, medical history, family history, physical activity levels, substance use (alcohol, tobacco, and drugs), and nutritional intake. Multiple psychosocial assessments evaluated perceived stress, reactions to racism, depression, sleep quality, anxiety, perceived social support, and pregnancy-related difficulties. Sonographic crown-rump length (CRL) was also recorded for all participants to confirm gestational age.

Study visit 2 included repeated measurements of blood pressure and weight, updates on physical activity level, substance exposure, and a psychological resilience questionnaire. Importantly, this visit incorporated a sonographic assessment of fetal biometry, cervical length, and uterine artery Doppler measurements. These parameters that have shown associations with preterm birth risk in previous research.

Study visit 3 collected additional clinical measurements and updated exposure assessments in the third trimester, a critical period for identifying imminent preterm birth risk. The sequential data collection process from preconception through post-delivery is detailed in Figure 1.

**Figure 1.**
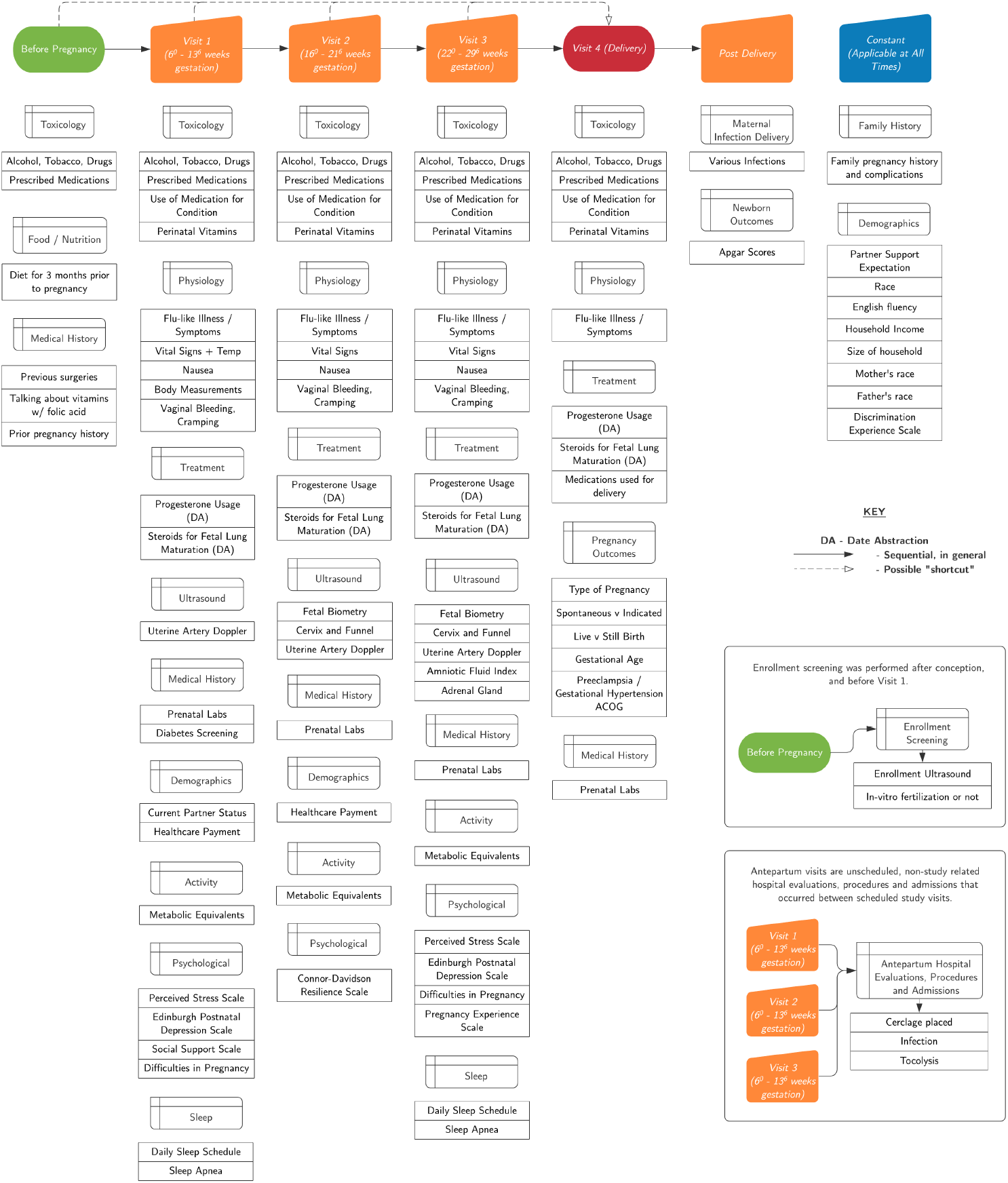
Timeline of Data Collection in Pregnancy and Postpartum Period. This figure outlines the sequential data collection process from preconception to post-delivery, detailing key assessments across different visits

The nuMoM2b dataset presents both challenges and opportunities for machine learning applications. It contains 10,426 features, of which 2,582 are obtained across the visit timepoints. Of these features, 1,425 are qualitative, while 9,001 are quantitative (numerical values). Missing values are an important consideration, with a mean missingness of 20.68% across all features, though the median missingness is 1.78%, indicating that most features have relatively complete data. This rich but complex dataset required extensive preprocessing to maximize its utility for prediction models, as described in detail by Anton et al.^47^

A key strength of this cohort is the availability of detailed information allowing differentiation between spontaneous and indicated preterm births, a critical distinction often lacking in previous prediction studies despite the fundamentally different biological mechanisms underlying these two preterm birth subtypes.

### Primary outcome for prediction

Our primary outcomes for prediction were indicated and spontaneous preterm births occurring before 37 completed weeks of gestation. We explicitly excluded fetal demise and stillbirth deliveries to focus specifically on live births. For classification purposes, we used clinically precise definitions to distinguish between the two major preterm birth subtypes:

*Spontaneous preterm birth (sPTLB)* was defined as delivery occurring after the spontaneous onset of preterm labor, premature rupture of the membranes (PROM), or fetal membrane prolapse.^46^ This category represents births initiated by physiological processes rather than medical intervention.

*Indicated preterm birth (iPTLB)* was defined as delivery following medical induction or cesarean section performed between 20^0^ and 36^6^ weeks gestation due to maternal or fetal complications that necessitated early delivery. These complications typically include conditions such as preeclampsia, intrauterine growth restriction, or maternal medical conditions requiring pregnancy termination.

In our analysis framework, we developed prediction models for three specific outcomes:

- Preterm live birth (PTLB): all preterm births regardless of subtype.
- Spontaneous preterm live birth (sPTLB): the subset of preterm births resulting from spontaneous processes.
- Indicated preterm live birth (iPTLB): the subset of preterm births resulting from medical intervention.

This stratified approach enabled us to examine whether predictive models perform differently across these clinically distinct preterm birth subtypes, potentially revealing different underlying biological mechanisms and risk factors.

### Preprocessing and Data Organization

The nuMoM2b dataset’s extensive feature set required substantial preprocessing to enable effective machine learning model development. To manage complexity and improve interpretability, we organized the features into 10 clinically relevant risk factor groups:

- **Treatment:** Variables related to treatments for already occurring preterm birth and medication administrations documented at the delivery visit.
- **Psychological:** Variables measuring the patient’s psychological state through validated clinical scales, including measures of depression, anxiety, stress, and resilience.
- **Medical History:** Variables relating to pre-existing conditions, long-term medical history, and various clinical tests performed on the mother and fetus/newborn during the study period.
- **Demographics:** Sociodemographic variables including race/ethnicity, education level, income, insurance status, and other social determinants of health.
- **Ultrasound:** Measurements from research and clinical ultrasounds, including cervical length, fetal biometry, and uterine artery Doppler studies.
- **Outcomes:** A metadata file containing variables that either contained privileged information for model training or classification labels for model evaluation.
- **Activity:** Variables quantifying maternal physical activity patterns, including exercise frequency, intensity, and duration.
- **Family History:** Variables capturing familial medical conditions and history that might influence pregnancy outcomes.
- **Diet:** Nutritional information covering the 3 months before pregnancy and throughout gestation.
- **Toxicology:** Variables related to substance exposure, including alcohol, tobacco, and drug use.

We conducted systematic reviews of the features of each risk factor group to establish evidence-based filtering rules, imputation strategies, and variable dependencies based on relevant medical literature. Our preprocessing prioritized the identification of high-level, general variables that could encompass related sub-questions asked in the questionnaires. This “filtering” process incorporated rules for variable summarization, scoring, scaling, and dependencies to reduce dimensionality while preserving clinically important information.

This comprehensive preprocessing approach reduced the feature set from over 10,000 to 398 variables selected for prediction model development. The average missingness across these selected features was approximately 37.67%. Figure 2 illustrates the distribution of missing values across the selected feature set.

**Figure 2.**
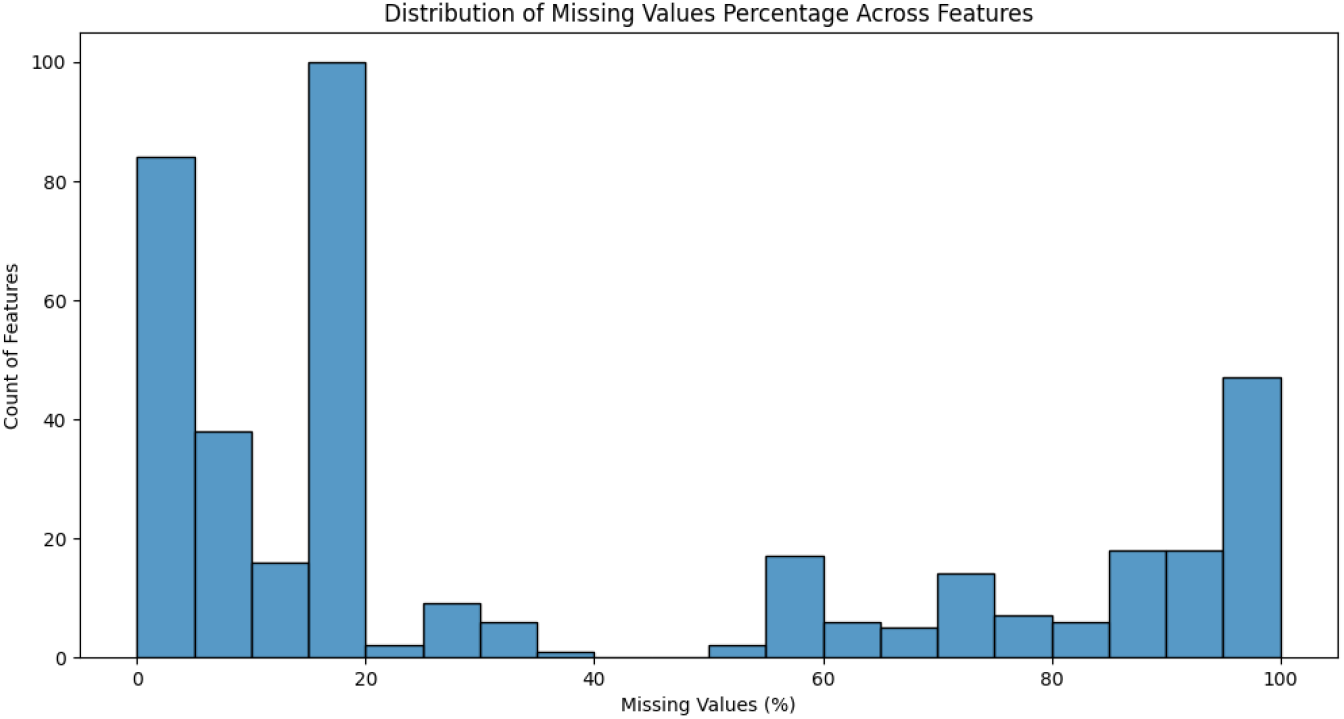
Histogram showing the distribution of missing values (%) across features in the dataset. The x-axis represents the percentage of missing values per feature, while the y-axis indicates the number of features with the corresponding missing value percentage. The plot highlights varying levels of missingness, with some features having minimal missing values and others having a high proportion of missing data.

To address the missing data challenge, we developed feature-specific imputation strategies rather than a one-size-fits-all approach. Each feature’s clinical significance, missingness pattern, and relationship with other variables informed its custom imputation method. This nuanced approach to missing data management helped preserve the integrity of the information while enabling comprehensive machine learning model development.

### Genetic Race Features

Understanding the role of genetic ancestry in preterm birth risk is critical, given established disparities across racial and ethnic groups. To move beyond self-reported race/ethnicity, we performed a rigorous genetic ancestry analysis on the nuMoM2b cohort. We determined the genetic ancestry of each participant using SNPweights v.2.1, leveraging approximately 40,000 ancestry informative markers available from the 1000 Genomes Consortium.

By applying an ancestry percentage threshold of *>* 50%, participants were stratified into five major population groups: South Asians (SAS), East Asians (EAS), Americans (AMR), Africans (AFR), and Europeans (EUR). Notably, we observed substantial genetic admixture within the cohort, prompting us to establish a sixth group (ADM) comprising subjects with no single ancestry group exceeding the 50% threshold. The distribution of participants across these ancestry groups is presented in Table 1.

**Table 1:** Breakdown of nuMoM2b subjects by genetic ancestry group.

| Ancestry | Number of Subjects |
| --- | --- |
| EUR | 6082 |
| AFR | 1425 |
| AMR | 846 |
| EAS | 323 |
| SAS | 122 |
| ADM | 891 |

We conducted a concordance analysis to assess the relationship between computed genetic ancestry and self-reported race. We found substantial overlap between genetic ancestry assignments and self-reported racial categories, with robust concordance between European ancestry and self-reported White race (5,108 individuals), African ancestry and self-reported Black race (1,162 individuals), and American ancestry and self-reported Hispanic ethnicity (675 individuals). Table 2 presents the full concordance matrix between self-reported race (columns) and computed genetic ancestry (rows).

**Table 2:** Concordance between self-reported race (columns) and pre-computed ancestry (rows)

|  | Asian | Black | Hispanic | Multiracial | White |
| --- | --- | --- | --- | --- | --- |
| AFR | 1 | **1162** | 94 | 86 | 6 |
| AMR | 0 | 2 | **675** | 9 | 104 |
| EAS | **239** | 0 | 6 | 15 | 2 |
| EUR | 6 | 12 | 376 | 155 | **5108** |
| ADM | 20 | 81 | **432** | 108 | 206 |
| SAS | **89** | 5 | 5 | 3 | 5 |

While genetic ancestry provides valuable biological insights, socioeconomic and environmental factors associated with self-identified race may also influence preterm birth risk through mechanisms not captured by genetics alone. Therefore, given the highly admixed nature of the cohort and to avoid overlooking important socioeconomic confounders, we ultimately used self-reported race for downstream analyses, including feature filtering, preprocessing, and imputation strategies. The primary correspondences between self-reported race and genetic ancestry that guided our approach were White - EUR, Black - AFR, Hispanic - AMR, Asian - EAS/SAS, and Multiracial - ADM.

This dual consideration of genetic ancestry and self-reported race allowed us to develop more nuanced prediction models to account for biological and social determinants of preterm birth risk. A detailed analysis of genetic associations of pregnancy traits for the nuMoM2b cohort is available through our prior works.^48,49^

### Odds Ratios Analysis

To identify potential risk factors for preterm birth and inform our feature selection process, we conducted a comprehensive odds ratio (OR) analysis on the nuMoM2b dataset. This univariate analysis served three key purposes: (1) to understand the dataset from a statistical perspective, (2) to provide a method for comparing features in our dataset with established risk factors from medical literature, and (3) to guide the feature selection process for our machine learning models.

We calculated odds ratios for a wide range of variables across different risk factor domains, including demographics, toxicology, physiology, physical activity, and psychological measures. For each variable, we computed odds ratios for three outcome categories: preterm live birth of less than 37 weeks gestational age (PTLB), spontaneous preterm live birth (sPTLB), and indicated preterm live birth (iPTLB). This stratified approach allowed us to identify risk factors that might be more strongly associated with one preterm birth subtype than the other.

Table c, in the results section, presents selected significant odds ratios from our analysis. The table highlights variables with statistically significant associations with preterm birth outcomes, along with their corresponding p-values. By computing separate odds ratios for each preterm birth subtype, we could observe differential risk patterns, thus identifying variables that exhibited stronger associations with either spontaneous or indicated preterm birth.

This odds ratio analysis provided valuable insights into which features, when analyzed in isolation, could be considered risk factors (OR *>* 1) or protective factors (OR *<* 1) for preterm birth. Additionally, the magnitude of these odds ratios offered a preliminary assessment of each feature’s potential predictive importance, which guided our subsequent machine learning approach.

The results of this analysis confirmed many known risk factors from the literature, such as maternal race, educational attainment, cervical length, and hypertensive disorders. Importantly, it also revealed distinct risk profiles for spontaneous versus indicated preterm birth, supporting our decision to develop separate prediction models for these clinically distinct outcomes. For instance, we observed that hypertensive disorders showed substantially stronger associations with indicated preterm birth, while cervical length measurements demonstrated stronger associations with spontaneous preterm birth.

Our comprehensive odds ratio analysis validated our approach against the existing medical literature but also provided data-driven insights for feature selection and model development in our machine learning pipeline.

### XGboost with Privileged Information (XGboost+)

Learning with Privileged Information (LUPI) represents an innovative paradigm in machine learning, introduced by Vapnik and Vashist.^50^ This approach leverages additional information available during model training that is inaccessible during evaluation or real-world deployment. Unlike conventional machine learning paradigms that rely solely on feature-label pairs, LUPI incorporates a third element: privileged information that enriches the learning process but cannot be utilized during inference.

In the context of preterm birth prediction, privileged information encompasses variables that would be unavailable when making predictions early in pregnancy but could inform the model’s understanding of risk patterns. Examples include the occurrence of adverse pregnancy outcomes (APOs) during later stages of pregnancy, post-delivery physiological information, and maternal outcomes recorded after birth. Additionally, clinical measurements from later prenatal visits can serve as privileged information when developing models that predict outcomes based on earlier visits.

### Theoretical Framework

The LUPI framework operates on triplets of information 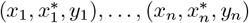, where *x*_*i*_ ∈ *X* represents standard features available during both training and deployment, 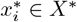 represents privileged information available only during training, and *y*_*i*_ ∈ {0, 1}represents the binary outcome (in our case, preterm birth status).

Vapnik and Vashist^50^ demonstrated that incorporating privileged information can substantially accelerate learning convergence from 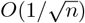. This theoretical improvement translates to significant practical benefits: a model with privileged information trained on 320 examples could potentially achieve the same performance as a conventional model trained on 100,000 examples 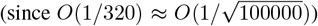. This represents a substantial efficiency gain with particular relevance in medical domains where large datasets may be difficult to obtain.

### XGboost+ Implementation

Drawing inspiration from Lopez-Paz et al.,^51^ we implemented the LUPI paradigm within the XGBoost framework using a generalized distillation approach. Our XGBoost+ model extends the standard XGBoost algorithm, which has proven highly effective for tabular data, to incorporate privileged information. This approach was first developed in our prior work on predicting proximal junctional kyphosis.^52^

In standard XGBoost with *K* additive learning rounds (also called boosting rounds), the predictive output is obtained by aggregating individual tree predictions:

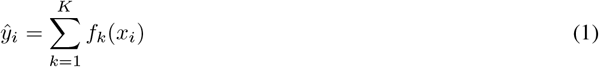

where *f*_*k*_ corresponds to a regression tree structure *q* with leaf weights *w*. The model is trained through a sequence of additive optimization steps. At iteration *t*, XGBoost aims to minimize the following objective function:

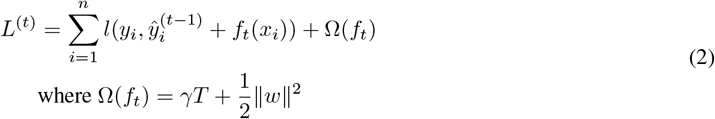

Here, *l* represents a twice-differentiable convex loss function that minimizes the difference between the target *y*_*i*_ and the prediction from the previous iteration 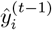 plus the current estimated residual error *f*_*t*_(*x*_*i*_). The regularization term Ω penalizes model complexity, with *T* representing the number of leaves in the tree and *w* representing the leaf weights.

To incorporate privileged information, we adapted the distillation framework proposed by Hinton et al.:^53^

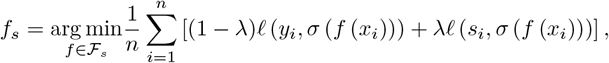

Where

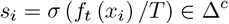

Lopez-Paz et al.^51^ adapted this framework for privileged information, which inspired our XGBoost+ approach. We modified the XGBoost objective function to incorporate privileged information as follows:

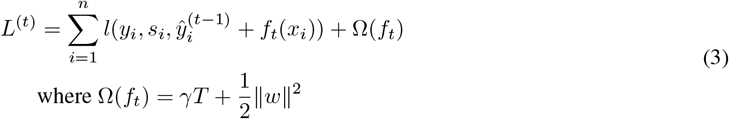

Using a second-order approximation, we compute:

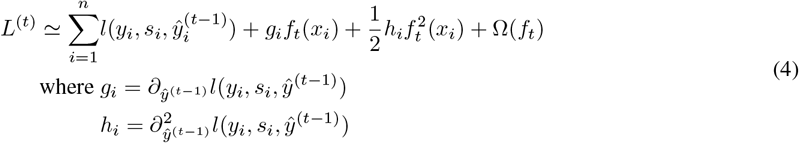

The privileged information is incorporated through 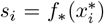, where 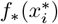 can be any classifier operating on the privileged feature space. The logistic loss function is updated as a weighted combination:

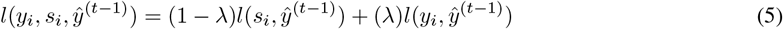

Where the individual loss terms are:

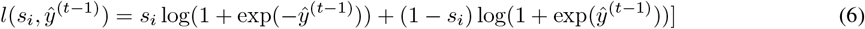

and

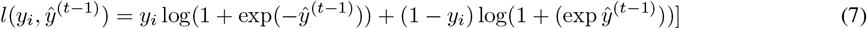

This formulation results in gradient statistics

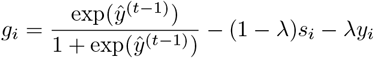

and

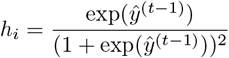

Importantly, by modifying only the loss function while retaining the core structure of the XGBoost algorithm, our XGBoost+ implementation preserves all the advantageous characteristics of standard XGBoost, including its weight quantile sketch for handling weighted data, sparsity-aware split finding for efficient handling of missing values, and parallel learning capabilities. For further technical details on our implementation, we refer readers to our previous work on proximal junctional kyphosis prediction.^52^

Through this integration of privileged information, XGBoost+ allows us to leverage data available during model development (such as outcomes from later pregnancy stages) to improve predictive performance at earlier time points, potentially enabling earlier clinical intervention for high-risk pregnancies.

### Sparsity-aware Split Finding

A significant challenge in medical datasets, including the nuMoM2b cohort, is the non-random nature of missing data. Unlike many datasets where missing values occur due to technical errors or collection oversights, medical data often contains meaningful patterns of missingness. For example, certain clinical tests may be selectively administered based on patient risk profiles, or patients may choose not to disclose specific health information. These patterns create systematic missingness that cannot be adequately addressed through conventional imputation techniques.

XGBoost provides a sophisticated approach to handling missing data through its sparsity-aware split-finding algorithm. This approach is particularly valuable for medical data analysis as it treats missingness as informative rather than problematic. When building each decision tree, XGBoost dynamically determines the optimal path for missing values at each split, effectively learning the most predictive way to handle missingness rather than relying on pre-specified imputation rules.

Specifically, for each node in a tree, XGBoost considers two potential directions for instances with missing values: they can either go with the left or right branches. The algorithm selects the direction that optimizes the overall objective function, treating “missing” as a separate value that can be learned from the data.

While we did implement feature-specific imputation strategies during preprocessing, as described earlier, these strategies inherently introduce assumptions and potential biases. For traditional machine learning algorithms such as Random Forest (RF), Support Vector Machine (SVM), Logistic Regression (LR), and Decision Tree (DT), these imputed values were necessary to enable model training. However, for our XGBoost-based models, we leveraged the native sparsity-aware capability, allowing the algorithm to learn directly from the patterns of missingness rather than relying solely on imputed values.

This approach was particularly important given the varied nature of missingness across the nuMoM2b dataset, with some features showing clinical or behavioral patterns in their missing values that could predict outcomes. By utilizing XGBoost’s sparsity-aware split finding, we maintained the integrity of the original data structure while maximizing predictive performance.

### Model building

We implemented a rigorous cross-validation modeling strategy to ensure reliable and generalizable prediction models. Our approach involved partitioning the data into training, validation, and test sets, with repeated trials to assess model stability and performance consistency.

### Stratified Cross-Validation

We employed a stratified cross-validation approach to account for the class imbalance inherent in preterm birth studies (approximately 10% preterm births in the general population). This ensured a proportional representation of both outcome classes (preterm and full-term) across all data partitions. For model development and evaluation, we created balanced training and test sets by undersampling the full-term cases while maintaining all preterm cases, resulting in a 1:1 ratio of preterm to full-term births in these sets.

### Hyperparameter Optimization

We conducted systematic hyperparameter optimization for each model type using the validation set. We employed Bayesian optimization^54^ to efficiently search the hyperparameter space.

For XGBoost and XGBoost+ models, we optimized key hyperparameters, including learning rate, maximum tree depth, minimum child weight, subsample ratio, column sample ratio, and the regularization terms alpha and lambda. For the XGBoost+ model, we additionally tuned the distillation parameter *λ* that balances the influence of true labels versus privileged information.

### Model Evaluation

We conducted 100 independent trials for each experiment to ensure robust performance assessment. For each trial, we:

1. Randomly partitioned the data into training (60%), validation (20%), and test (20%) sets using stratified sampling
2. Trained models on the training set with hyperparameters optimized on the validation set
3. Evaluated final model performance on the held-out test set

We assessed model performance using the area under the receiver operating characteristic curve (AUC-ROC), providing a threshold-independent discriminative ability measure. Additionally, we calculated sensitivity, specificity, and precision at an operating threshold of 0.5 to assess clinical utility. All performance metrics were averaged across the 100 trials to provide robust estimates of model performance.

### Comparative Analysis

We implemented multiple machine learning algorithms to benchmark against our XGBoost+ approach, including:

1. Logistic Regression (LR): A traditional statistical approach widely used in clinical prediction.
2. Support Vector Machine (SVM): A robust classifier for finding optimal decision boundaries.
3. Decision Tree (DT): A simple, interpretable model that forms the building blocks of ensemble methods.
4. Random Forest (RF): An ensemble of decision trees that mitigates overfitting through bagging.
5. XGBoost: The standard gradient boosting implementation without privileged information.
6. XGBoost+: Our proposed model incorporating privileged information. This comparative framework allowed us to assess the relative advantages of each approach for preterm birth prediction and precisely evaluate the benefit of incorporating privileged information through our XGBoost+ methodology.

### Software packages

All data processing and model development were implemented in Python 3.7, with extensive use of scientific computing libraries to ensure reproducibility and scalability. Our implementation leveraged the following key packages:

- **XGBoost (version 3.0.0)**: For implementing both standard XGBoost models and our XGBoost+ extension with privileged information. We utilized XGBoost’s Python API for model training, prediction, and feature importance analysis.
- **scikit-learn (version 1.6.1)**: For implementing comparative machine learning models (Logistic Regression, SVM, Decision Trees, Random Forest), cross-validation procedures, and performance evaluation metrics.
- **imbalanced-learn (version 0.13.0)**: For handling class imbalance through undersampling and oversampling techniques, ensuring balanced training datasets while preserving the integrity of minority class examples.
- **pandas (version 2.2.3)** and **NumPy (version 2.2.2)**: For efficient data manipulation, feature engineering, and numerical computations.
- **BayesianOptimization (version 2.0.3)**: For hyperparameter tuning using Bayesian optimization techniques, which efficiently explore hyperparameter spaces to identify optimal model configurations.
- **matplotlib (version 3.10.0)** and **seaborn (version 0.11.1)**: For data visualization and results presentation, including ROC curves, feature importance plots, and performance comparison charts.
- **SHAP (version 0.47.1)**: For model interpretability and explanation through SHapley Additive exPlanations, providing insights into feature contributions to individual predictions.

To facilitate reproducibility and transparency, the complete codebase for this study, including data preprocessing, model development, and evaluation, is available in the PRAISE-Lab GitHub repository: https://github.com/PRAISE-Lab-Repository/PTB_Prediction.git. This repository includes detailed documentation on data structures, preprocessing pipelines, model configurations, and evaluation procedures to enable replication of our findings and extension to other research contexts.

For researchers interested in applying our XGBoost+ methodology to other clinical prediction problems, we have also provided a standalone implementation of the XGBoost+ algorithm with documentation and usage examples in the repository. This implementation is designed to be generalizable to supervised learning tasks where privileged information is available during training but not during deployment or inference.

## Results

### Dataset

Our analysis focused on live births from pregnancies of nulliparous women in the nuMoM2b cohort. To ensure clarity of outcomes, we systematically excluded records of stillbirths and fetal demises, as illustrated in Figure 3. Additionally, we excluded from the analysis any patients who delivered during the chronological period corresponding to specific visit time points, ensuring that our predictive models for each trimester contained only data from pregnancies that were still ongoing at that point.

**Figure 3.**
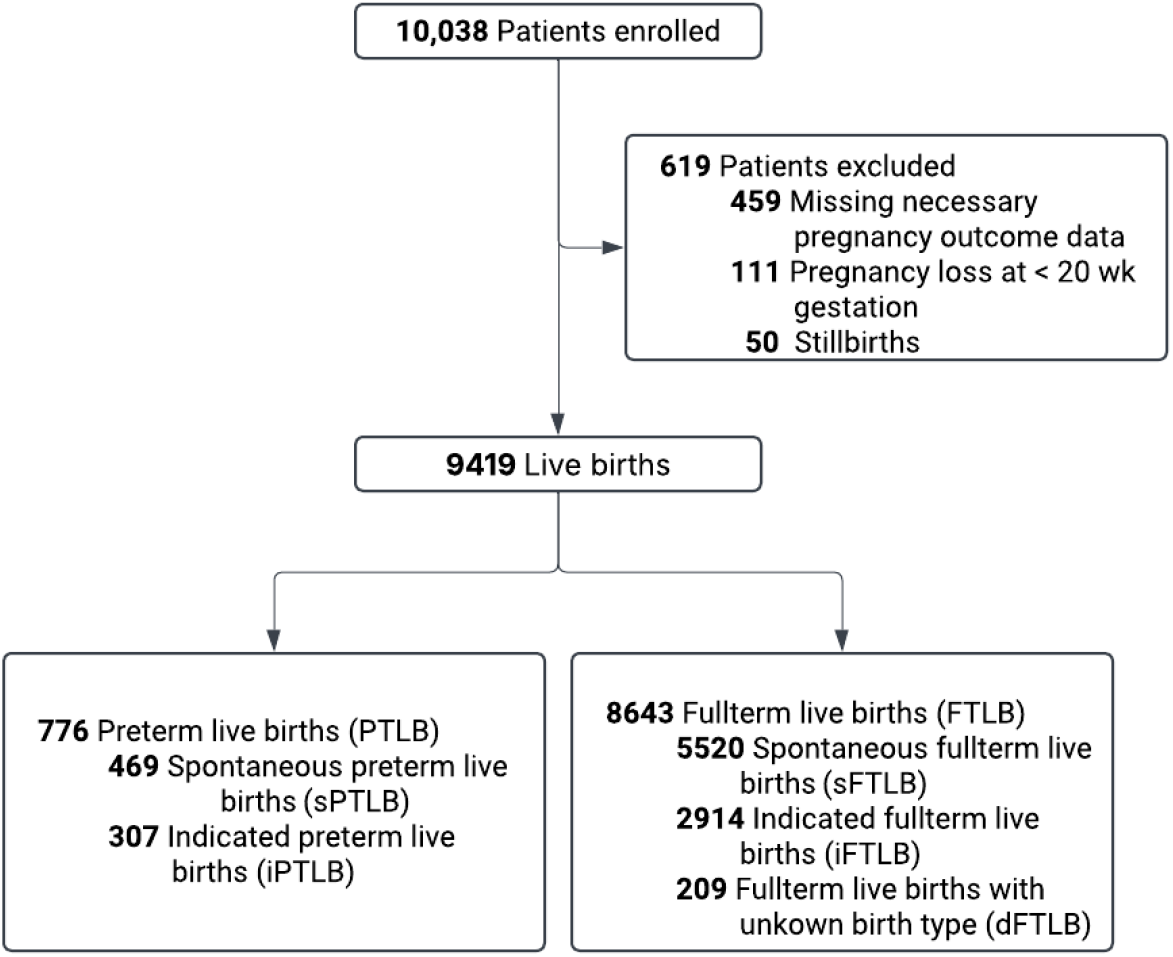
Flowchart of patient enrollment and classification for the preterm birth prediction model. A total of 10,038 patients were enrolled, with 619 exclusions due to missing data, early pregnancy loss, or stillbirths. The final dataset included 9,419 live births, categorized into preterm (PTLB) and full-term (FTLB) births, with further sub-classifications based on spontaneous and indicated birth types.

Figure 3 depicts the comprehensive patient flow through the study, beginning with 10,038 enrolled participants and documenting the systematic exclusion of 619 cases due to missing necessary pregnancy outcome data (459 cases), pregnancy loss before 20 weeks gestation (111 cases), or stillbirths (50 cases). The final analytical cohort comprised 9,419 live births, which we further stratified into preterm (776) and full-term (8,643) categories. We then applied clinical definitions to sub-classify the preterm births into spontaneous (469) and indicated (307) subtypes and the full-term births into spontaneous (5,520), indicated (2,914), and unspecified birth type (209) categories.

Table 3 provides a detailed summary of the distribution of birth outcomes by type and study visit. The classification includes spontaneous full-term live birth (sFTLB), indicated full-term live birth (iFTLB), deliveries with unspecified birth type (dFTLB), indicated preterm live birth (iPTLB), and spontaneous preterm live birth (sPTLB). We also document the number of features incorporated into each visit’s predictive model, reflecting the accumulation of clinical data as the pregnancy progressed: 241 features at visit 1, 312 at visit 2, and 398 at visit 3.

**Table 3:**
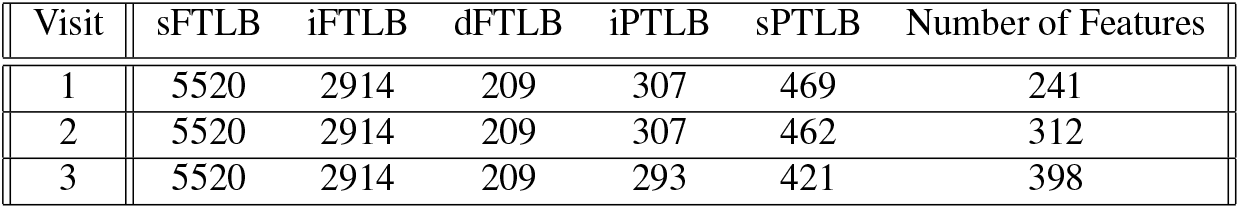
Summary of birth outcomes and predictive features by study visit. The table shows the distribution of birth types (sFTLB: spontaneous full-term live birth; iFTLB: indicated full-term live birth; dFTLB: delivery with unspecified birth type; iPTLB: indicated preterm live birth; sPTLB: spontaneous preterm live birth) and the number of features incorporated into predictive models for each visit timepoint.

| Visit | sFTLB | iFTLB | dFTLB | iPTLB | sPTLB | Number of Features |
| --- | --- | --- | --- | --- | --- | --- |
| 1 | 5520 | 2914 | 209 | 307 | 469 | 241 |
| 2 | 5520 | 2914 | 209 | 307 | 462 | 312 |
| 3 | 5520 | 2914 | 209 | 293 | 421 | 398 |

**Table 4:** Odds ratios and p-values for top risk factors associated with preterm birth in the NuMoM2b dataset. The table presents the odds ratios for all preterm live births (PTLB), indicated preterm live births (iPTLB), and spontaneous preterm live births (sPTLB) across various demographic, clinical, and socioeconomic factors.

| Risk Factor | Odds Ratio |  |  | p-value |  |  |
| --- | --- | --- | --- | --- | --- | --- |
|  | PTLB | iPTLB | sPTLB | PTLB | iPTLB | sPTLB |
| **Maternal Age at V1 (Ref. Group 20-34)** |  |  |  |  |  |  |
| < 20 | 1.69 | 1.87 | 1.60 | < 0.001 | < 0.001 | < 0.001 |
| > 34 | 1.55 | 1.53 | 1.54 | < 0.001 | 0.02 | 0.01 |
| **Gravidity (Ref. 1)** |  |  |  |  |  |  |
| 2 | 1.06 | 1.12 | 1.03 | 0.52 | 0.48 | 0.80 |
| ≥ 3 | 1.51 | 2.12 | 1.14 | 0.01 | < 0.001 | 0.47 |
| **Maternal Race / Ethnicity (Ref. Non-Hispanic White)** |  |  |  |  |  |  |
| Non-Hispanic Black | 1.80 | 2.22 | 1.54 | < 0.001 | < 0.001 | < 0.001 |
| Hispanic | 1.08 | 1.21 | 1.02 | 0.51 | 0.30 | 0.89 |
| Asian | 0.79 | 1.09 | 0.70 | 0.31 | 0.84 | 0.23 |
| Native Hawaiian | 1.64 | 0.00 | 3.08 | 0.32 | 0.62 | 0.06 |
| Other of Missing | 0.57 | 0.86 | 0.43 | 0.77 | 1.00 | 0.72 |
| Multiracial | 1.43 | 1.43 | 1.44 | 0.04 | 0.25 | 0.12 |
| **Education Status Attained (Ref. Completed college)** |  |  |  |  |  |  |
| Less than HS grad | 2.05 | 1.86 | 2.16 | < 0.001 | 0.01 | < 0.001 |
| HS grad or GED | 1.79 | 1.60 | 1.92 | < 0.001 | 0.03 | < 0.001 |
| Some college | 1.55 | 1.61 | 1.48 | < 0.001 | 0.01 | 0.01 |
| Assoc/Tech degree | 1.43 | 1.28 | 1.53 | 0.01 | 0.25 | 0.02 |
| Degree work beyond college | 0.94 | 0.82 | 1.02 | 0.60 | 0.33 | 0.94 |
| **Income & Household Size (Ref. > 200% of fed poverty level )** |  |  |  |  |  |  |
| 100-200% of fed poverty level | 1.25 | 1.34 | 1.19 | 0.08 | 0.14 | 0.28 |
| < 100% of fed poverty level | 1.52 | 1.91 | 1.30 | < 0.001 | < 0.001 | 0.08 |
| **Cervical Length in mm (Ref. Group 32-47)** |  |  |  |  |  |  |
| ≤ 25 | 3.70 | 2.72 | 4.36 | < 0.001 | < 0.001 | < 0.001 |
| 25-32 | 2.02 | 1.66 | 2.29 | < 0.001 | 0.01 | < 0.001 |
| > 47 | 0.75 | 0.78 | 0.72 | 0.03 | 0.29 | 0.07 |
| **Method of Paying for Healthcare (Ref. Commercial)** |  |  |  |  |  |  |
| Govt insurance | 1.68 | 1.82 | 1.59 | < 0.001 | < 0.001 | < 0.001 |
| Military insurance | 1.46 | 1.33 | 1.53 | 0.32 | 0.66 | 0.35 |
| Other | 2.06 | 0.82 | 2.84 | 0.03 | 1.00 | < 0.001 |
| Personal | 1.02 | 1.64 | 0.70 | 0.90 | 0.15 | 0.43 |
| **Hypertensive Conditions (Ref. No hypertensive disorder)** |  |  |  |  |  |  |
| Eclampsia | 8.33 | 36.55 | < 0.001 | 0.05 | 0.01 | 1.00 |
| Superimposed preeclampsia | 22.78 | 39.16 | 3.67 | < 0.001 | < 0.001 | 0.29 |
| Severe preeclampsia | 11.30 | 21.51 | 2.95 | < 0.001 | < 0.001 | < 0.001 |
| Mild preeclampsia | 1.66 | 2.75 | 1.19 | 0.03 | < 0.001 | 0.57 |
| New onset antepartum HTN | 1.40 | 1.99 | 1.29 | 0.02 | < 0.001 | 0.23 |
| New onset intrapartum/postpartum HTN | 0.78 | 0.59 | 0.88 | 0.09 | 0.12 | 0.51 |
| **Diabetes (Ref. Group No pre-gestational DM or GDM)** |  |  |  |  |  |  |
| Gestational diabetes | 1.95 | 1.16 | 2.98 | < 0.001 | 0.53 | < 0.001 |
| Pre-gestational diabetes | 6.44 | 5.27 | 7.98 | < 0.001 | < 0.001 | < 0.001 |
| **Tobacco (Ref. No)** |  |  |  |  |  |  |
| Yes | 1.95 | 1.85 | 2.04 | < 0.001 | 0.06 | < 0.001 |
| **Depression Summary During V3 (Ref. Group 26-30)** |  |  |  |  |  |  |
| ≤ 25 | 2.51 | 2.42 | 2.52 | < 0.001 | 0.01 | < 0.001 |
| 31-35 | 0.83 | 0.70 | 0.95 | 0.14 | 0.08 | 0.73 |
| > 35 | 0.90 | 0.68 | 1.09 | 0.39 | 0.06 | 0.67 |
| **Stress Summary During V3 (Ref. 21-28)** |  |  |  |  |  |  |
| ≤ 20 | 0.92 | 0.87 | 0.96 | 0.38 | 0.35 | 0.74 |
| 29-35 | 1.12 | 1.11 | 1.10 | 0.41 | 0.61 | 0.59 |
| 21-28 | 1.00 | 1.00 | 1.00 | 1.00 | 1.00 | 1.00 |
| > 35 | 2.40 | 2.11 | 2.58 | < 0.001 | 0.02 | < 0.001 |

### Exploratory Data Analysis

#### Differential Risk Profiles for Preterm Birth Subtypes

Our univariate analysis of the nuMoM2b dataset revealed distinct risk profiles for indicated and spontaneous preterm births, providing compelling evidence for fundamentally different pathophysiological mechanisms underlying these two clinical entities. Table c presents the odds ratios for key risk factors stratified by preterm birth subtype.

Among cardiovascular and metabolic risk factors, hypertensive disorders demonstrated the most striking subtype-specific associations. Superimposed preeclampsia exhibited an overall odds ratio of 22.78 (p*<*0.001) for preterm birth.

Still, when analyzed by subtype, the association was dramatically stronger for indicated preterm birth (OR=39.16, p*<*0.001) compared to spontaneous preterm birth (OR=3.67, p=0.29). Similarly, severe preeclampsia showed an overall odds ratio of 11.30 (p*<*0.001), with substantially higher risk for indicated (OR=21.51, p*<*0.001) versus spontaneous (OR=2.95, p*<*0.001) preterm birth. This pronounced difference in cardiovascular risk provides strong clinical evidence for distinct etiological pathways between the two preterm birth subtypes. It explains the superior predictive performance of our models for indicated preterm birth.

Cervical structure emerged as another risk factor with subtype-specific associations but with the opposite pattern. Short cervical length (≤ 25 mm) demonstrated a stronger association with spontaneous preterm birth (OR=4.36, *p <* 0.001) compared to indicated preterm birth (OR=2.72, p*<*0.001). This finding aligns with the established mechanical role of cervical integrity in maintaining pregnancy and preventing spontaneous preterm labor. Miller et al.^55^ previously reported an odds ratio of 5.90 (CI: 3.72-9.36) for cervical length ≤ 25*mm* and preterm birth, which is higher than but consistent with our findings.

Pre-gestational diabetes was significantly associated with both preterm birth subtypes, though with a stronger relationship to spontaneous (OR=7.98, p*<*0.001) than indicated (OR=5.27, p¡0.001) preterm birth. This finding suggests potential metabolic influences on spontaneous labor mechanisms warrant further investigation.

Psychosocial factors demonstrated important associations with preterm birth risk. Depression, measured at visit 3 with scores ≤ 25 (indicating more depression), showed similar associations with both indicated (OR=2.42, p=0.01) and spontaneous (OR=2.52, p*<*0.001) preterm birth. High-stress scores (*>*35) at visit 3 demonstrated a slightly stronger association with spontaneous (OR=2.58, p*<*0.001) than indicated (OR=2.11, p=0.02) preterm birth, suggesting that psychological factors can influence spontaneous labor mechanisms more directly.

Tobacco use within the previous month showed a dose-response relationship with advancing gestational age, with odds ratios increasing from 1.87 at visit 1 to 1.95 at visit 4 (p*<*0.001 for both). When analyzed by subtype, tobacco use was more strongly associated with spontaneous (OR=2.04, p*<*0.001) than indicated (OR=1.85, p=0.06) preterm birth, consistent with its known effects on placental function and fetal growth.

We also identified protective factors, most notably physical activity. Higher metabolic equivalent task (METs) scores, particularly at visits 2 and 3, were associated with reduced preterm birth risk. For example, MET scores *>*12 at visit 3, indicative of high physical activity, showed a strong protective effect (OR=0.41, p*<*0.001). Among women with MET scores *>*12 at visit 3, only 27 delivered preterm compared to 606 who delivered at term. This finding is consistent with previous research by Catov et al.,^56^ who reported an odds ratio of 0.76 (CI: 0.61-0.95) for high leisure-time physical activity throughout pregnancy.

Demographic factors demonstrated important associations, particularly with respect to race/ethnicity and socioeconomic indicators. Non-Hispanic Black race showed an identical odds ratio (1.80, p*<*0.001) to that reported in previous literature, but with a stronger association with indicated (OR=2.22, p*<*0.001) than spontaneous (OR=1.54, p*<*0.001) preterm birth. Educational attainment below the high school level showed a gradient effect, with stronger associations for spontaneous (OR=2.16, p*<*0.001) than indicated (OR=1.86, p=0.01) preterm birth. Conversely, income below the federal poverty level demonstrated a stronger relationship with indicated (OR=1.91, p*<*0.001) than spontaneous (OR=1.30, p=0.08) preterm birth.

This comprehensive odds ratio analysis reveals fundamental differences in risk profiles between preterm birth sub-types. Indicated preterm birth appears more strongly associated with vascular/metabolic disorders, higher gravidity, and certain socioeconomic factors. In contrast, spontaneous preterm birth demonstrates stronger connections to cervical structure, gestational diabetes, tobacco use, and psychological stress. These distinct risk profiles support our approach of developing separate prediction models for each preterm birth subtype and highlight the importance of targeted surveillance and management strategies based on specific risk profiles.

#### Feature Selection and Data Leakage Mitigation

To ensure robust model development, we conducted systematic evaluations to identify and mitigate potential sources of data leakage. These are features that might artificially inflate model performance through information not realistically available at the time of prediction. Through iterative testing and feature importance analysis, we identified several potential leakage sources:

1. **Exercise indicators:** We found that metabolic equivalent task (METs) scores could introduce leakage, as clinicians often recommend reduced physical activity for women already identified as high-risk for preterm birth. This clinical intervention creates a reverse causality pattern that could artificially improve prediction.
2. **Hypertensive conditions:** While strong predictors of indicated preterm birth, diagnosed conditions like preeclampsia often directly trigger medical intervention for delivery, creating near-perfect prediction that does not represent true prospective value.
3. **Missing diagnostic tests:** We observed missingness patterns in third-trimester Group B Streptococcus (GBS) culture results were highly informative. Preterm deliveries often lack these results not because of random missingness but because the test was never performed due to early delivery, creating systematic missingness patterns that artificially improve prediction.

For each identified source of potential leakage, we implemented specific mitigation strategies, including feature transformation, careful temporal alignment of predictors and outcomes, and developing models that explicitly account for the clinical decision-making processes that generate the data. This systematic approach to leakage mitigation ensures that our reported model performance reflects true predictive capacity rather than data collection or clinical practice artifacts.

### Preterm Birth Prediction Models

#### Comparative Model Performance

We evaluated multiple machine learning approaches for preterm birth prediction, with results summarized in Table 5. All models demonstrated modest performance at visit 1 (early pregnancy), with progressive improvement as additional data became available at visits 2 and 3, reflecting the accumulation of clinically relevant information throughout pregnancy.

**Table 5:** Comparison of model performance for predicting preterm birth across pregnancy time points. Values represent mean AUC *±* standard deviation across 100 independent trials. DT: Decision Tree; RF: Random Forest; SVM: Support Vector Machine; LR: Logistic Regression. Bold values indicate the best-performing model at each visit.

| Visit | DT | RF | SVM | LR | XGBoost | XGBoost+ |
| --- | --- | --- | --- | --- | --- | --- |
| 1 | 0.54 ± 0.03 | 0.59 ± 0.03 | 0.59 ± 0.03 | 0.59 ± 0.04 | 0.60 ± 0.03 | **0.63 ± 0.03** |
| 2 | 0.58 ± 0.03 | 0.63 ± 0.03 | 0.63 ± 0.03 | 0.61 ± 0.04 | 0.64 ± 0.03 | **0.66 ± 0.03** |
| 3 | 0.61 ± 0.03 | 0.68 ± 0.03 | 0.65 ± 0.04 | 0.67 ± 0.02 | 0.70 ± 0.03 | **0.72 ± 0.03** |

The Decision Tree (DT) classifier consistently demonstrated the lowest performance, with AUC values ranging from 0.54 at visit 1 (barely above random classification) to 0.61 at visit 3. This limited performance likely reflects the inability of simple tree structures to capture the complex, multifactorial nature of preterm birth risk.

More sophisticated approaches, such as Random Forest (RF), Support Vector Machine (SVM), and Logistic Regression (LR), achieved moderate performance improvements, with visit 3 AUC values of 0.68, 0.65, and 0.67, respectively. The gradient boosting approach implemented in XGBoost further improved performance, reaching an AUC of 0.70 at visit 3.

Most notably, our novel XGBoost+ model, which incorporates privileged information, consistently outperformed all other approaches across all visit time points. At visit 1, when clinical data are most limited, XGBoost+ achieved an AUC of 0.63, representing a three percentage point improvement over standard XGBoost. By visit 3, XGBoost+ reached an AUC of 0.72, demonstrating the added value of privileged information even with a rich feature set. This consistent 2-3 percentage point improvement across all time points validates the theoretical advantages of the privileged information framework in the context of preterm birth prediction.

#### Differential Performance for Preterm Birth Subtypes

A more detailed analysis of the visit 3 model performance, presented in Table 6, reveals important differences in predictive capability for different preterm birth subtypes. Figure 4 illustrates these differences through receiver operating characteristic (ROC) curves for XGBoost and XGBoost+ models.

**Table 6:** Detailed performance analysis of XGBoost and XGBoost+ models at visit 3, evaluated at a classification threshold of 0.5. Results are stratified by preterm birth type, demonstrating the differential predictability of indicated versus spontaneous preterm birth. Bold values indicate the better model for each metric within each category.

| Category | Method | Specificity | Sensitivity | Precision | AUC |
| --- | --- | --- | --- | --- | --- |
| All Preterm Births | XGBoost | 0.75 | **0.54** | 0.68 | 0.70 |
|  | XGBoost+ | **0.89** | 0.38 | **0.78** | **0.72** |
| Indicated Preterm Births | XGBoost | 0.68 | **0.69** | 0.68 | 0.74 |
|  | XGBoost+ | **0.84** | 0.60 | **0.79** | **0.78** |
| Spontaneous Preterm Births | XGBoost | 0.68 | **0.58** | 0.65 | 0.67 |
|  | XGBoost+ | **0.87** | 0.34 | **0.74** | **0.68** |

**Figure 4.**
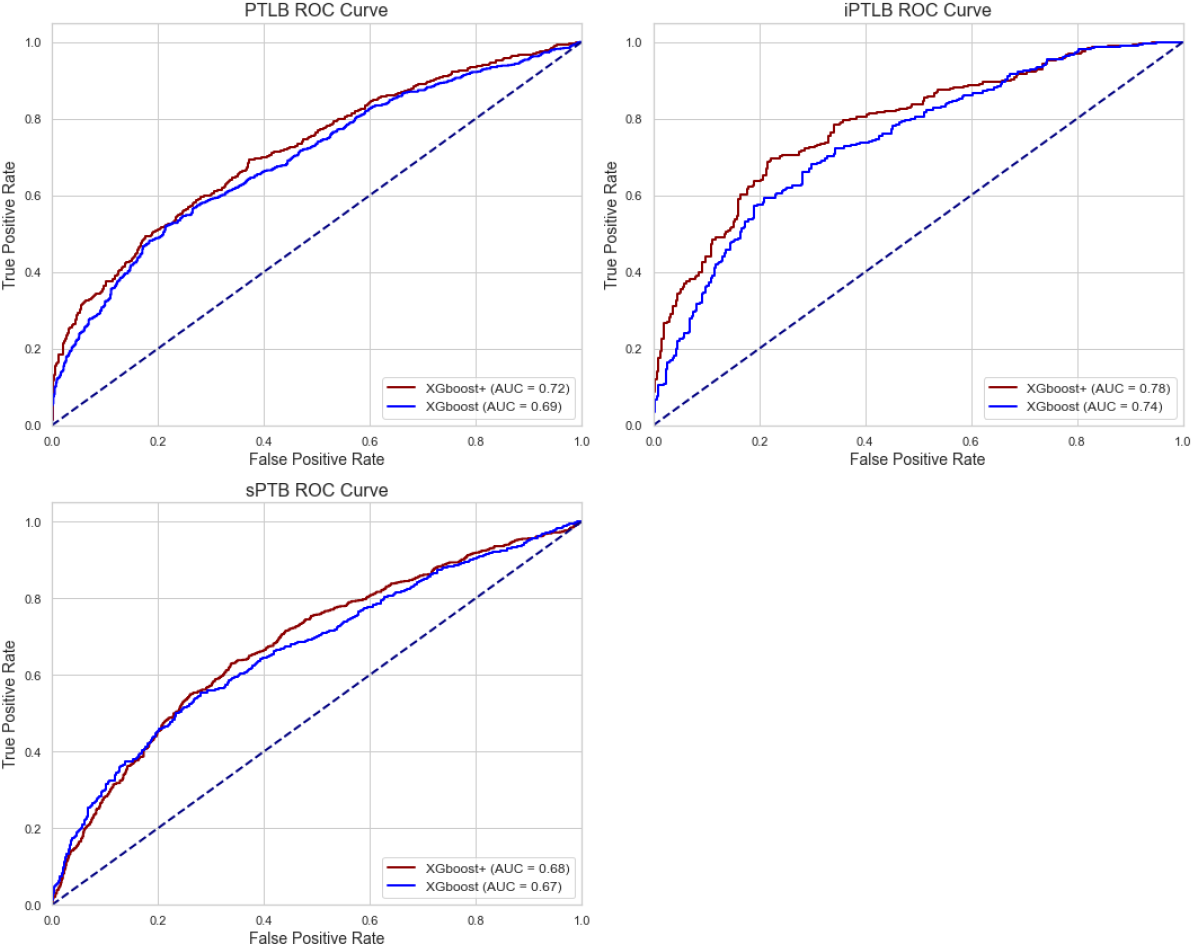
Receiver Operating Characteristic (ROC) curves comparing XGBoost and XGBoost+ models at visit 3 for predicting all preterm births (PTLB), indicated preterm births (iPTLB), and spontaneous preterm births (sPTLB). The curves illustrate the superior discrimination capability of XGBoost+ for all outcomes, with the largest improvement observed for indicated preterm birth prediction.

For overall preterm birth prediction (PTLB), XGBoost+ demonstrated higher specificity (0.89 vs. 0.75) and precision (0.78 vs. 0.68) than standard XGBoost when evaluated at a classification threshold of 0.5. This improved specificity and precision came at the cost of reduced sensitivity (0.38 vs. 0.54), reflecting a trade-off that favors more confident positive predictions. The overall AUC improvement from 0.70 to 0.72 confirms that XGBoost+ provides better discrimination across all possible thresholds.

We observed a striking difference in model performance when examining preterm birth subtypes. For indicated preterm birth (iPTLB), both models achieved substantially higher AUC values (0.74 for XGBoost and 0.78 for XG-Boost+) than for spontaneous preterm birth (0.67 for XGBoost and 0.68 for XGBoost+). This difference highlights the greater predictability of medically indicated preterm deliveries, which often follow from identifiable physiological complications.

Notably, the performance gain from incorporating privileged information was substantially larger for indicated preterm birth (AUC improvement of 0.04) than spontaneous preterm birth (AUC improvement of 0.01). This differential improvement suggests that the privileged information-particularly data related to hypertensive conditions and other pregnancy complications-provides more relevant signals for predicting indicated preterm birth than for spontaneous preterm birth. This finding aligns with clinical understanding, as indicated preterm births are often precipitated by conditions such as preeclampsia and other hypertensive disorders that may manifest subtly earlier in pregnancy.

#### Feature Importance Analysis

To elucidate our models’ key drivers of predictive performance, we employed SHapley Additive exPlanations (SHAP)^57^ to quantify and visualize feature contributions in the XGBoost+ model at visit 3. SHAP plots show how each feature impacts the prediction of a model. In a beeswarm SHAP plot:

1. Features are listed vertically in order of importance.
2. The x-axis shows SHAP values. Negative values decrease the prediction, while positive values increase it.
3. Each dot represents an instance/patient.
4. The color indicates the value of the feature (red = high, blue = low).
5. The horizontal spread shows the range and distribution of the impact.
6. The vertical position within a feature row is only for visibility (jitter).

Figure 5 and 6 presents the SHAP summary plot for the top 15 features, revealing distinct patterns of predictive importance.

**Figure 5.**
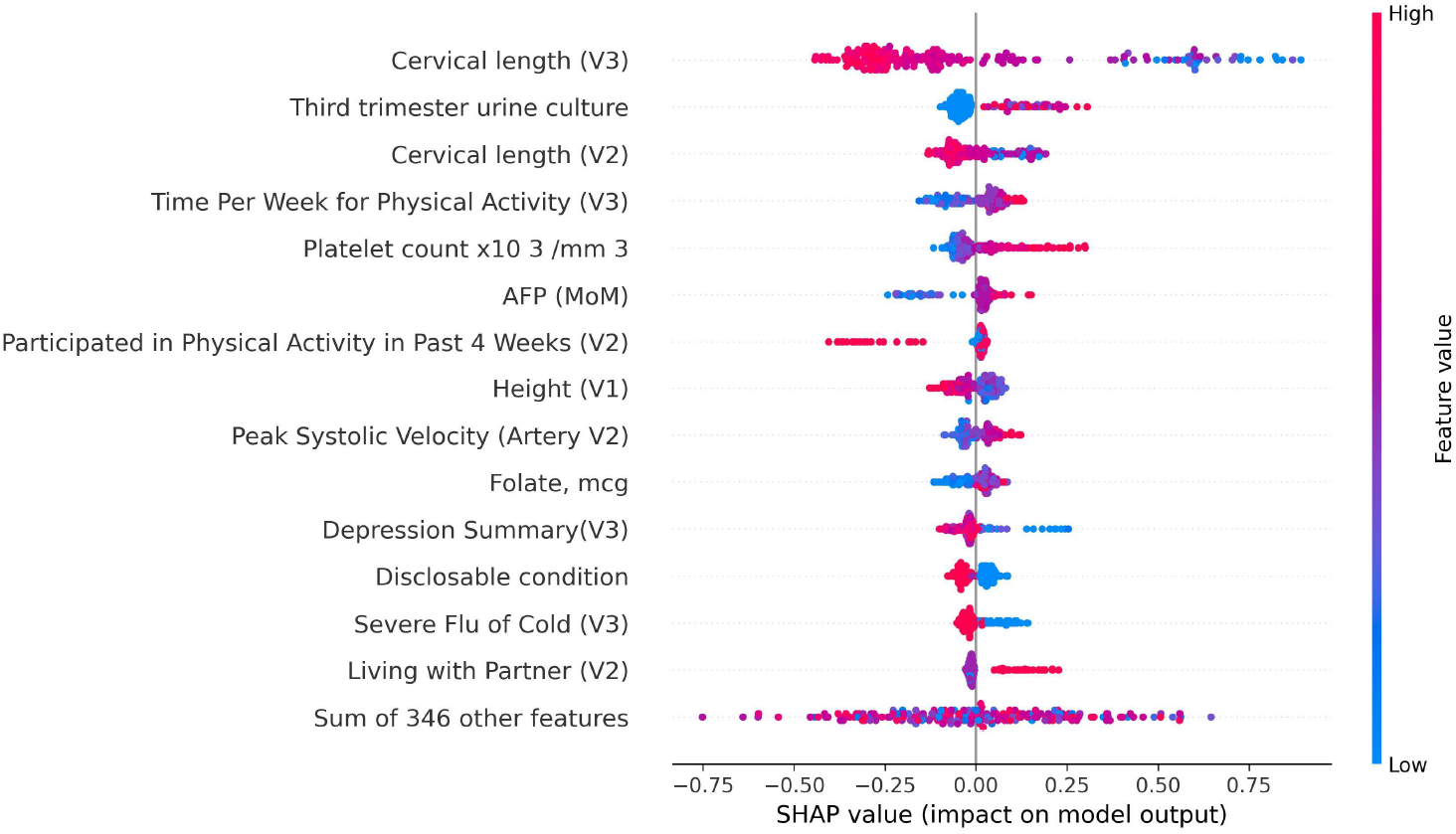
SHAP (SHapley Additive exPlanations) plot displaying the impact of the top 15 features on prediction output for the XGBoost+ model at visit 3 for predicting spontaneous preterm birth.

**Figure 6.**
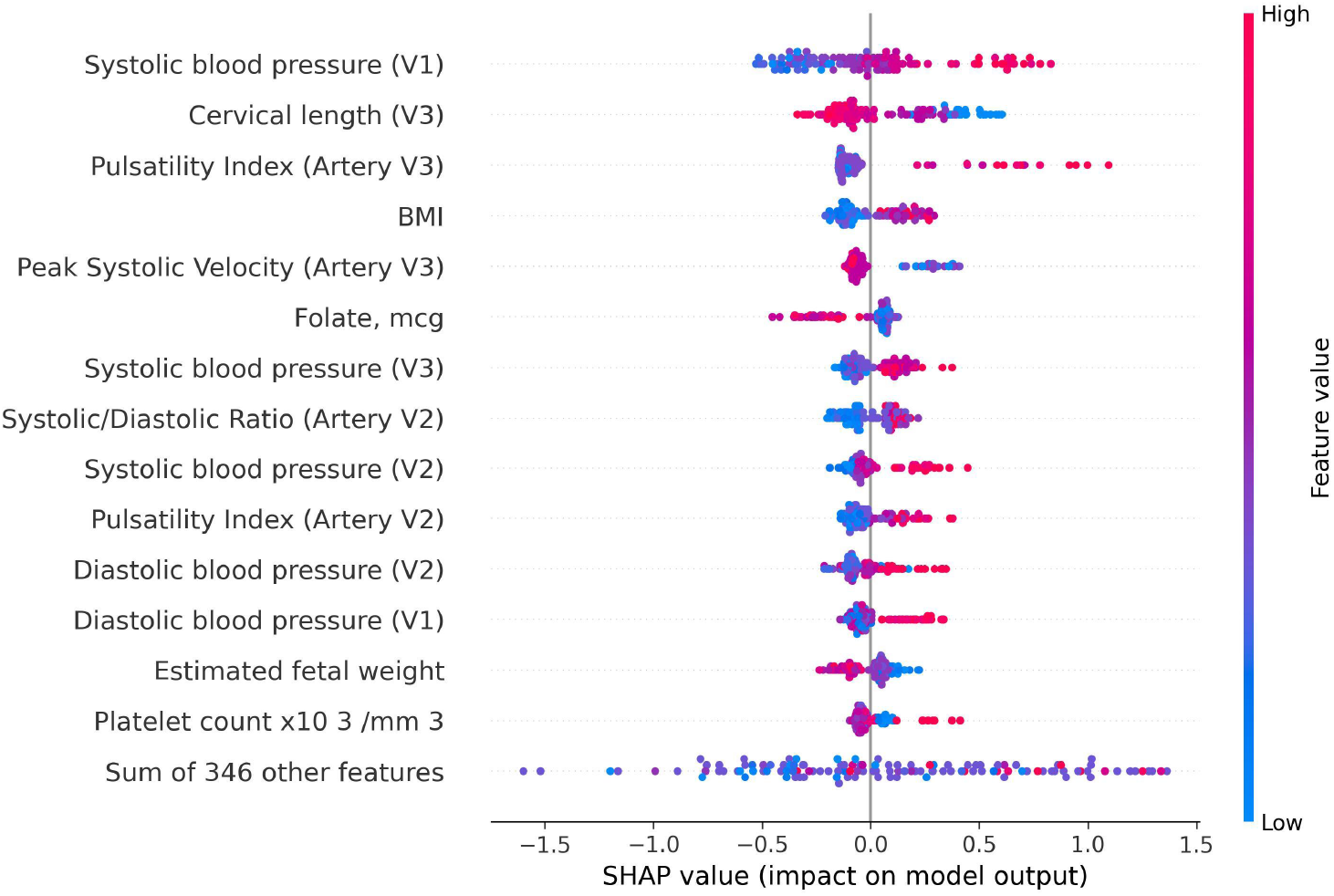
SHAP (SHapley Additive exPlanations) plot displaying the impact of the top 15 features on prediction output for the XGBoost+ model at visit 3 for predicting indicated preterm birth.

Cervical length measured at visit 3 emerged as the single most influential predictor, with shorter lengths strongly associated with an increased risk of preterm birth, particularly for spontaneous preterm birth. The impacts of visit 3 cervical length is much higher in spontaneous subtype versus indicated which demonstrated by the wider distribution of SHAP values. This finding aligns with the established mechanical role of the cervix in maintaining pregnancy and the clinical use of cervical length as a screening tool for the risk of preterm birth.

Third-trimester urinary tract infection status, as indicated by urine culture results, demonstrated significant predictive value, primarily for spontaneous preterm birth. Positive cultures were associated with increased risk, consistent with the known relationship between maternal infection and inflammatory processes that can trigger preterm labor.

Hematological parameters, particularly platelet count, ranked among the top predictors. Higher platelet counts, which can indicate the development of blood clots or, paradoxically, bleeding. It may signal underlying inflammatory or placental disorders associated with spontaneous preterm birth.

Cardiovascular measures, including systolic and diastolic blood pressure at visit 3, demonstrated substantial predictive importance, primarily for indicated preterm birth. These hemodynamic parameters serve as early indicators of developing hypertensive disorders that frequently necessitates medically indicated preterm delivery. The prediction of preeclampsia is discussed in our previous work,^58^ which was selected as the winner of the NICHD’s Decoding Maternal Morbidity Data Challenge.^59^ Along with cardiovascular measure, indicator such as high maternal BMI and abnormal uterine artery pulsatility index can also be indicators of preeclampsia.

Estimated fetal weight is directly related to fetal growth restriction, which is another major cause of indicated preterm birth.

Nutritional factors, including folate intake, exhibited meaningful predictive importance. Lower folate levels were associated with increased preterm birth risk for both subtype, consistent with the established role of folate in placental development and function.

This feature importance analysis reveals the most predictive individual factors and illuminates the distinct patho-physiological pathways underlying indicated versus spontaneous preterm birth. The predominance of vascular, and fetal pathology are among the top predictors for indicated preterm birth contrasts with the importance of cervical parameters, infection markers, and stress indicators for spontaneous preterm birth, further supporting the fundamental biological differences between these clinical entities.

## Discussion

Our comprehensive investigation of preterm birth prediction in nulliparous women reveals critical insights into the distinct nature of spontaneous and indicated preterm birth subtypes and their differential predictability using machine learning approaches. By leveraging the extensive nuMoM2b dataset with over 10,000 features collected across multiple time points in pregnancy, we have developed predictive models that demonstrate practical clinical utility and illuminate fundamental biological differences between preterm birth subtypes.

### Differential Predictability of Preterm Birth Subtypes

The most striking finding of our study is the markedly higher predictive performance for indicated preterm birth compared to spontaneous preterm birth. Our XGBoost+ model achieved an AUC of 0.78 for indicated preterm birth but only 0.68 for spontaneous preterm birth at visit 3. This substantial difference in predictability reflects the distinct etiological pathways underlying these two clinical entities.

Indicated preterm birth typically results from identifiable maternal complications that necessitate medical intervention for delivery. These complications-particularly hypertensive disorders such as preeclampsia-often develop through progressive physiological dysregulation that manifests in measurable clinical parameters well before the decision to deliver. Blood pressure measurements, platelet counts, and other biomarkers captured in routine clinical care provide early signals of these developing complications. Our predictive models effectively capture these progressive patterns, resulting in superior performance for indicated preterm birth prediction.

In contrast, spontaneous preterm birth remains remarkably challenging to predict, particularly in early pregnancy. Unlike indicated preterm birth, which follows recognizable pathophysiological trajectories, spontaneous preterm birth appears to result from multiple, often subclinical processes that may not manifest in routine clinical measurements until labor onset is imminent. The complex interplay of inflammatory, neuroendocrine, mechanical, and vascular factors that potentially trigger spontaneous preterm labor remains poorly captured by conventional clinical data.

Even with the rich feature set in the nuMoM2b cohort and our sophisticated machine learning approach, spontaneous preterm birth prediction achieved only modest performance. This underscores a fundamental limitation: spontaneous preterm birth may be inherently less predictable from standard clinical data, suggesting that critical biological signals are either not being measured or occur too proximally to the event to enable timely intervention.

### Value of Privileged Information Framework

Our novel application of the Learning with Privileged Information paradigm through the XGBoost+ model demonstrates meaningful performance improvements across all preterm birth categories. By incorporating information available during model training but not during deployment, XGBoost+ achieved consistent AUC improvements of 2-4 percentage points compared to conventional XGBoost.

Notably, the performance gain was substantially larger for indicated preterm birth (AUC improvement of 0.04) than for spontaneous preterm birth (AUC improvement of 0.01). This differential benefit aligns with our understanding of the pathophysiological processes involved. The privileged information-particularly data related to pregnancy complications and outcomes that manifest later in gestation-provides more relevant signals for the progressive conditions that lead to indicated preterm birth than for the potentially more acute or subclinical processes driving spontaneous preterm birth.

The practical implication of this finding extends beyond the specific models developed in this study. It suggests that even with access to perfect knowledge of all pregnancy outcomes and complications, our ability to predict spontaneous preterm birth from clinical data alone would remain limited. This fundamental constraint highlights the need for novel biomarkers and monitoring approaches that capture the biological processes more proximal to spontaneous labor onset.

### Clinical and Biological Insights from Model Features

Feature importance analysis through SHAP values provides valuable insights into the biological mechanisms underlying preterm birth subtypes. The dominance of vascular and metabolic parameters among top predictors for indicated preterm birth-ncluding blood pressure measurements, platelet counts, and abdominal circumference-aligns with the established role of maternal cardiovascular and metabolic dysfunction in conditions like preeclampsia that frequently necessitate medically indicated delivery.

For spontaneous preterm birth, cervical length emerged as the single most important predictor, consistent with its mechanical role in maintaining pregnancy. The significance of infection markers, such as urinary tract infection status, reflects the established relationship between maternal infection, inflammation, and the triggering of preterm labor. Psychological factors, including stress and depression scores, demonstrated meaningful predictive importance for spontaneous preterm birth, supporting the growing recognition of psychoneuroimmunological pathways in spontaneous labor initiation.

The odds ratio analysis further reinforces these distinct risk profiles. The dramatically higher odds ratios for hypertensive disorders with indicated versus spontaneous preterm birth (e.g., OR=39.16 vs. OR=3.67 for superimposed preeclampsia) represent striking evidence of fundamentally different causal pathways. Similarly, the stronger associations of cervical length, psychological stress, and tobacco use with spontaneous preterm birth highlight the multifactorial nature of this outcome.

These distinct risk profiles improve our understanding of preterm birth etiology and suggest potential targets for subtype-specific interventions. The high predictability of indicated preterm birth related to hypertensive disorders points to the value of enhanced monitoring and management of cardiovascular risk factors. In contrast, the challenges in spontaneous preterm birth prediction suggest the need for more comprehensive approaches addressing cervical integrity, infection prevention, and psychological well-being.

### Methodological Strengths and Limitations

Several methodological aspects strengthen the validity and generalizability of our findings. The nuMoM2b cohort-a prospective, multi-site study designed to investigate nulliparous pregnancy outcomes-provides a rich, well-characterized dataset with standardized data collection across diverse populations. The rigorous cross-validation approach with multiple independent trials ensures robust performance estimation. At the same time, our systematic evaluation and mitigation of potential data leakage sources enhance the clinical relevance of our findings.

The application of multiple machine learning approaches, from simple decision trees to sophisticated ensemble methods, allows for comprehensive performance comparison and confidence in the superior results achieved by our XGBoost+ model. The SHAP-based feature importance analysis provides interpretable insights that connect statistical patterns to biological mechanisms, enhancing the clinical utility of our models.

Despite these strengths, several limitations warrant consideration. First, while the nuMoM2b cohort is large and diverse, it remains a selected population that may not fully represent all pregnant individuals, particularly those with limited access to prenatal care. Second, our analysis relies on features collected through conventional clinical assessment, potentially missing important biological signals that are not routinely measured. Third, the binary classification of preterm birth at *<*37 weeks gestation simplifies a complex phenotype that includes varying degrees of prematurity with potentially different predictive patterns.

Additionally, while our models demonstrate good discriminative ability, their clinical implementation would require careful consideration of the appropriate operating threshold to balance sensitivity and specificity based on the specific clinical context and intervention cost-benefit considerations. Finally, although we have identified predictive patterns, these associations do not necessarily imply causality and the biological interpretation of complex machine learning models remains challenging.

### Implications for Future Research and Clinical Practice

Our findings have important implications for research priorities and clinical practice in preterm birth prevention. The superior predictability of indicated preterm birth suggests that focused attention on early identification and management of hypertensive disorders and other conditions that lead to medically indicated delivery could yield substantial benefits. Enhanced monitoring protocols for high-risk individuals, particularly those with elevated cardiovascular risk factors identified by our models, could enable earlier intervention and potentially delay delivery to reduce prematurity-related complications.

For spontaneous preterm birth, the modest predictive performance from clinical data highlights the critical need for novel approaches focused on more proximal biological processes. Future research should prioritize developing and validating biomarkers that capture the inflammatory, neuroendocrine, and mechanical pathways involved in spontaneous labor initiation. Potential directions include:

1. High-resolution cervical imaging beyond simple length measurement, potentially incorporating texture analysis, elastography, or other advanced techniques to assess cervical remodeling processes better.
2. Longitudinal assessment of vaginal microbiome composition and function, which has shown promising associations with spontaneous preterm birth risk.
3. Novel inflammatory biomarkers that more specifically reflect the maternal-fetal interface and may provide earlier signals of the inflammatory cascade that can trigger preterm labor.
4. Integration of genetic and epigenetic markers with environmental exposures to capture gene-environment interactions that influence preterm birth risk.
5. Continuous monitoring approaches that track physiological parameters throughout pregnancy to identify subtle pattern changes that precede clinical manifestations.

From a methodological perspective, our successful application of the privileged information framework suggests potential value in applying this approach to other clinical prediction problems where information asymmetry exists between model development and deployment. The consistent performance improvements achieved by XGBoost+ demonstrate that leveraging additional information during training, even if not available during inference, can enhance model learning and generalization.

## Conclusion

Our extensive investigation of preterm birth prediction in nulliparous women has revealed critical insights into the fundamental differences between indicated and spontaneous preterm birth subtypes. Through comprehensive analysis of the nuMoM2b cohort and application of advanced machine learning techniques, we have demonstrated that indicated preterm birth can be predicted with good accuracy (AUC 0.78) using our novel XGBoost+ approach. In contrast, spontaneous preterm birth remains stubbornly difficult to predict from clinical data alone (AUC 0.68).

Indicated preterm birth, primarily driven by hypertensive disorders and other maternal complications, follows more predictable pathophysiological trajectories that manifest in measurable clinical parameters. Our models effectively capture these progressive patterns, offering the potential for early identification and targeted management of high-risk pregnancies. The successful prediction of indicated preterm birth represents a significant advancement that could enable timely interventions to optimize outcomes when early delivery becomes necessary.

In stark contrast, spontaneous preterm birth prediction remains a substantial challenge, even with comprehensive clinical data and sophisticated machine learning approaches. The modest predictive performance achieved for spontaneous preterm birth suggests fundamental limitations in the information content of conventional clinical measurements for capturing the complex, multifactorial biological processes that trigger spontaneous labor. This finding underscores the likelihood that spontaneous preterm birth may be driven by processes that are either not captured in routine clinical assessment or occur too proximally to the event to enable prediction from early pregnancy data.

Our novel XGBoost+ methodology, which incorporates privileged information available during model training but not during deployment, consistently improved predictive performance across all outcomes. The differential benefit observed-larger for indicated than spontaneous preterm birth-further reinforces these clinical entities’ distinct nature and underlying biological mechanisms.

The distinct risk profiles identified through our odds ratio analysis and feature importance evaluation provide valuable insights for targeted preventive strategies. Focusing on cardiovascular risk factor management may yield significant benefits for indicated preterm birth. For spontaneous preterm birth, comprehensive approaches addressing cervical integrity, infection prevention, and psychological well-being may be necessary, though likely insufficient, without additional biological monitoring.

Future research should prioritize the development of novel biomarkers and monitoring approaches that capture the proximal biological processes leading to spontaneous preterm labor. Particularly promising directions include advanced cervical imaging, vaginal microbiome assessment, and longitudinal inflammatory profiling that may detect subtle changes preceding clinical manifestations of preterm labor. Since sPTB arise from multiple biological pathways, another promising direction^60^ we are investigating is to leverage recent advances in generative algorithms to up-sample data from the various pathways. We can then generate more accurate models for the sPTB class.

In conclusion, our work demonstrates the potential and limitations of clinical data for predicting preterm birth. While indicated preterm birth shows encouragingly high predictability, the persistent challenge of spontaneous preterm birth prediction suggests that this complex phenotype likely requires fundamental advances in understanding and measuring the biological processes that initiate spontaneous labor. By clearly delineating these differential predictive patterns, our research provides a framework for more targeted approaches to preterm birth prevention based on subtype-specific risk profiles and biological mechanisms.

## Data Availability

All data produced are available online at NICHD Dash: Nulliparous Pregnancy Outcomes Study: Monitoring Mothers-to-Be (nuMoM2b)

https://dash.nichd.nih.gov/study/226675

## Acknowledgments

This study was supported by grant funding from the Eunice Kennedy Shriver National Institute of Child Health and Human Development (NICHD) grant number R01LM013327. The funder played no role in study design, data collection, analysis and interpretation of data, or the writing of this manuscript.

All research design, data analysis, scientific interpretations and conclusions were conceived and directed entirely by the authors of this manuscript. The authors acknowledge the use of Claude, an Anthropic AI assistant. The AI tool served solely to enhance the clarity and readability of the manuscript while preserving the authors’ original scientific contributions and intellectual content. The primary prompt used was “Can you revise this section to enhance the grammar and flow?”

## Data Availability

The datasets generated and analyzed during the current study were obtained from the NIH Data and Specimen Hub (DASH). Access restrictions apply to these data in accordance with data use agreements and participant privacy protections. While the raw data are not publicly available due to these restrictions, researchers may request access through formal application to the NIH DASH repository. Qualified researchers can obtain the data by submitting a request to the repository administrators and securing appropriate institutional permissions. The authors will facilitate data access requests when possible, subject to compliance with all applicable regulations and the original data use agreement terms.

## Ethical Approval

This study, titled “SCH: Prediction of Preterm Birth in Nulliparous Women,” was conducted with full ethical oversight and approval. The research protocol underwent comprehensive review and received approval from both the Columbia University Human Subjects Institutional Review Board and the City University of New York (CUNY) Institutional Review Board. All research activities were performed in accordance with relevant guidelines and regulations for human subjects research. Informed consent was obtained from all participants prior to their inclusion in the study.

